# Predicting symptomatic response to prokinetic treatment using Gastric Alimetry

**DOI:** 10.1101/2025.01.30.25321436

**Authors:** Chris Varghese, Sibylle Van Hove, Gabriel Schamberg, Billy Wu, Nooriyah Poonawala, Mikaela Law, Nicky Dachs, Gen Johnston, India Fitt, Daphne Foong, Henry P. Parkman, Thomas Abell, Vincent Ho, Stefan Calder, Armen A Gharibans, Christopher N Andrews, Gregory O’Grady

## Abstract

**Background:** Chronic neurogastroduodenal disorders are challenging to manage, with therapy often initiated on a trial and error basis. Prokinetics play a significant role in management, but responses are variable and have been associated with adverse events, impacting widespread use. We investigated whether body surface gastric mapping (BSGM) biomarkers (using Gastric Alimetry**^®^**) could inform patient selection for prokinetic therapy.

**Methods:** Patients with chronic gastroduodenal symptoms taking oral prokinetic, regardless of gastric emptying status, were prospectively recruited and underwent BSGM (30 m baseline, 482 kcal standardised meal, 4 h postprandial recording) whilst off prokinetic. Patients were followed up with daily symptom diaries. A subset was compared to matched patients not taking prokinetics. Prokinetic responders were defined based on symptom improvement greater than a minimum clinically important difference methodology.

**Key Results:** 42 patients (88% female; median age 36; median BMI 26) taking prokinetics were analysed. Prokinetic prescribing, compared to matched patients, was independent of BSGM metrics (p>0.15). In patients on existing prokinetics (withheld for BSGM), lower amplitudes predicted reduced symptom burden, whereas low rhythm stability predicted a worse symptom burden (p<0.05). In prokinetic-naive patients (i.e. started on a prokinetic during the study), a lower postprandial amplitude predicted responders (mean 37.5±10.6 uV in responders [n=5] vs mean 54.8±6.6 uV among non-responders [n=3], p=0.047).

**Conclusions:** Gastric Alimetry biomarkers may help in the prediction of prokinetic response in patients with chronic gastroduodenal symptoms. Lower post-prandial amplitudes, indicating a reduced meal response, appear to predict benefit, whilst impaired rhythm stability predicted poorer therapeutic response.

## Introduction

Chronic neurogastroduodenal disorders, including chronic nausea and vomiting syndromes and gastroparesis, are challenging to manage, with therapy often initiated on a trial and error basis. Prokinetics remain a therapeutic mainstay in gastroparesis,^1,2^ but also variably demonstrate benefit with normal gastric emptying such as in functional dyspepsia.^3^ Further, the severity of gastric emptying delays do not correlate well to overall symptom responses to prokinetics.^4,5^

Metoclopramide is the most widely used prokinetic in the United States, while domperidone is commonly used in other parts of the world. Both agents function primarily through dopamine D_2_ receptor antagonism, enhancing gastric motility and providing anti-nausea properties. Prucalopride, a selective serotonin 5-HT4 receptor agonist, is also widely available and is approved for chronic idiopathic constipation and sometimes used off-label for gastroparesis.^6^ However, long term use of several prokinetics may be associated with significant side effects. D_2_ antagonists may carry risks of extrapyramidal symptoms (oral metoclopramide in particular) and a propensity to induce cardiac arrhythmias.^7–9^ Additionally, responses to prokinetic agents are generally highly variable, with no clear prognostic biomarkers available to guide optimal patient selection.^10,11^

Recently, Gastric Alimetry® has emerged as a non-invasive test of gastric function, offering new biomarkers to guide therapeutic management.^12^ In this prospective observational study, we applied Gastric Alimetry body surface gastric mapping (BSGM) to investigate whether gastric myoelectrical function could inform the selection of patients most likely to respond to prokinetic therapy.

## Methods

This was a prospective observational cohort study with a pre-specified and published protocol approved by the Auckland Health Research Ethics Committee (AHREC; ref AH1130).^13^ All patients provided informed consent. This study is reported in accordance with the STROBE guidelines.^14^

### Selection criteria

Patients with chronic gastroduodenal symptoms clinically diagnosed as functional dyspepsia (FD), chronic nausea and vomiting syndromes (CNVS), or gastroparesis were enrolled. Exclusion criteria included age <18 years, history of skin allergies or a history of extreme sensitivity to cosmetics or lotions, and vulnerable groups such as prisoners, individuals known to have cognitive impairment or institutionalised individuals. Patients were excluded if they had a pyloric procedure (pyloric botulinum toxin, pyloroplasty or gastric peroral endoscopic myotomy) between the baseline and on-prokinetic symptom evaluations. Additionally, patients who received pyloric botulinum toxin within 180 days prior to the baseline symptom evaluation were also excluded so as to mitigate confounding treatment effects.

### Gastric Alimetry protocol and interpretation

All participants underwent BSGM testing (30 m fasting, 482kcal standardised meal, and 4 h postprandial recording with simultaneous symptom profiling). All patients abstained from medications altering gastric motility for 48 h (including prokinetics). Gastric Alimetry spectral phenotyping followed established consensus criteria based on overall 4 h post-prandial data.^15^ In brief, neuromuscular phenotype was defined by an overall Gastric Alimetry Rhythm Index (GA-RI) <0.25, and/or overall low amplitudes <22 μV,^16^ and high-frequency phenotype was defined as a Principal Gastric Frequency >3.35 cycles per minute (cpm), with the remaining classified as having a normal spectral phenotype on the basis of otherwise normal rhythm, amplitude and frequency metrics.

### Outcomes

Patients were asked to complete an FDA-approved gastroduodenal symptom daily diary for 1 week at repeated intervals, commencing on days 0, 30, 90 and 180 after testing.^17^ Patients were classified as prokinetic responders if their change in weekly mean total symptom score (TSS) between baseline and a post-treatment daily diary was greater than the minimum clinically important difference (MCID); defined as the standard error of the mean (SEM) change in TSS whereby a positive difference indicates a reduction in TSS.

### Data analysis

Prokinetic use was the exposure of interest, and all prokinetics were considered under a unified group in this first BSGM study, regardless of their mechanism of action. Two primary analyses were performed to capture all participants on prokinetics during the study and follow-up period.

Firstly, all participants who were on existing prokinetic therapy prior to index Gastric Alimetry testing (always withheld 48h+ prior to testing) were compared to matched controls (i.e., symptomatic patients not on prokinetics). Patients on prokinetics were matched for age, sex, body mass index (BMI), neuromodulator use, and anti-emetic use, via propensity scores using a k-nearest-neighbours algorithm. It was ensured no patients had pyloric procedures within 180 days of study start date to confound treatment effects. Thereafter, a general linear model was constructed to evaluate the interaction between BSGM metrics, phenotypes, and prokinetic use. These results were presented with β coefficients and 95% confidence errors calculated based on robust standard errors in conjunction with their adjusted marginal means.

Secondly, a change analysis of all patients that had symptoms measured whilst off prokinetics for at least 48 h were analysed, with comparison to daily symptom diary results after starting a prokinetic. This analysis included patients that were previously on prokinetic therapies but were asked to withhold these medications prior to Gastric Alimetry testing; an off-drug baseline measurement of symptoms and physiological data were captured. This group also included a second subset of patients that were previously prokinetic naive, and subsequently started a prokinetic during follow-up, and who were evaluated in an independent subgroup analysis. Again the MCID of the change in symptom scores were used to define responders. The MCID is reported as the SEM of two TSS scores (mean difference ± standard deviation). Independent samples t tests and Chi square tests were used to compare efficacy of matching, and also Gastric Alimetry biomarkers and phenotypes between groups. Paired samples t tests were used to compare baseline TSS to on-prokinetic TSS. All analyses were performed in Python v3.9.7 and R v.4.4.2 (R Foundation for Statistical Computing, Vienna, Austria).

## Results

42 patients with complete follow-up symptom diaries were analysed (88% female; median age 36 [IQR 25-53] years; and median BMI 26 [IQR 20-31]). Cohort demographics across each analytical cohort are summarised in **Table 1**. Prokinetics prescribed included domperidone (n=22), prucalopride (n=9), metoclopramide (n=7), erythromycin (n=2), pyridostigmine (n=1) and mosapride (n=1). Of the 42 patients on a prokinetic during the study period, 34 were using prokinetics prior to Gastric Alimetry testing, and 8 were prokinetic naive and started using a prokinetic during follow-up.

**Table 1:** Demographics across each analysed cohort.

|  | Matched Analysis |  |  | Change Analysis |  |  |
| --- | --- | --- | --- | --- | --- | --- |
|  | On prokinetics<br>(n=32) | Matched patients not on prokinetics<br>(n=32) | *p* | Overall (n=42) | Subgroup (n=8) | *p* |
| **Demographics** |  |  |  |  |  |  |
| Sex (Female) | 27 (84.38%) | 23 (71.88%) | 0.36 | 37 (88.10%) | 8 (100.00%) | 0.70 |
| Age | 36.00 (26.00-53.25) | 39.00 (26.75-56.00) | 0.43 | 35.50 (24.50-52.75) | 28.50 (23.25-44.25) | 0.61 |
| BMI | 22.20 (19.20-29.52) | 24.85 (21.65-27.85) | 0.95 | 25.45 (19.45-31.18) | 33.05 (27.17-39.52) | 0.11 |
| Test duration | 4:40 (4:38-4:42) | 4:40 (4:38-4:40) | 0.42 | 4:40 (4:38-4:41) | 4:40 (4:39-4:40) | 0.68 |
| Percent artifact | 19.20 (10.40-25.12) | 23.50 (15.40-32.32) | 0.20 | 18.35 (10.20-24.60) | 18.00 (11.12-21.12) | 0.79 |
| **Ethnicity** |  |  |  |  |  |  |
| Asian | 1 (3.12%) | 3 (9.38%) | 0.61 | 1 (2.38%) | 0 (0%) | 1.00 |
| European / White | 25 (78.12%) | 22 (68.75%) | 0.57 | 29 (69.05%) | 4 (50.00%) | 0.53 |
| Other / Not Reported | 6 (18.75%) | 7 (21.88%) | 1.00 | 12 (28.57%) | 4 (50.00%) | 0.44 |
| **Medication** |  |  |  |  |  |  |
| Neuromodulator | 19 (59.38%) | 14 (43.75%) | 0.32 | 23 (54.76%) | 3 (37.50%) | 0.61 |
| Antiemetic | 13 (40.62%) | 10 (31.25%) | 0.60 | 16 (38.10%) | 2 (25.00%) | 0.76 |
| Opioid | 6 (18.75%) | 4 (12.50%) | 0.73 | 9 (21.43%) | 3 (37.50%) | 0.60 |
| Laxative | 11 (34.38%) | 3 (9.38%) | 0.03 | 14 (33.33%) | 2 (25.00%) | 0.96 |
| **BSGM Symptoms** |  |  |  |  |  |  |
| Total symptom burden | 21.15 (9.47-29.05) | 13.85 (7.05-21.98) | 0.24 | 18.55 (9.03-27.00) | 13.70 (7.62-18.38) | 0.47 |
| Bloating | 3.35 (1.30-5.60) | 2.50 (0.28-4.95) | 0.30 | 3.35 (1.52-5.45) | 2.65 (1.65-4.58) | 0.60 |
| Early satiety | 4.00 (0.00-8.00) | 4.50 (0.00-6.25) | 0.54 | 4.50 (0.00-7.75) | 4.50 (0.00-7.00) | 0.80 |
| Heartburn | 0.35 (0.00-3.52) | 0.00 (0.00-0.97) | 0.06 | 0.30 (0.00-3.33) | 0.45 (0.00-1.95) | 0.81 |
| Excessive fullness | 3.60 (0.97-6.33) | 3.00 (0.78-5.12) | 0.39 | 3.60 (1.00-5.50) | 2.00 (1.12-4.30) | 0.38 |
| Nausea | 1.55 (0.40-4.33) | 1.45 (0.00-4.10) | 0.59 | 1.70 (0.40-4.38) | 1.15 (0.30-2.58) | 0.50 |
| Upper gut pain | 3.10 (1.05-4.00) | 2.35 (0.28-3.82) | 0.67 | 2.95 (0.83-4.07) | 0.80 (0.33-2.33) | 0.24 |
| Stomach burn | 1.00 (0.00-2.80) | 0.00 (0.00-1.32) | 0.32 | 0.55 (0.00-2.70) | 0.15 (0.00-2.38) | 0.95 |

### Matched analysis: effect of prokinetics on symptoms

32 patients on prokinetic therapy that had completed a minimum of 30-day follow-up were matched to controls based on age, sex, BMI, and co-prescription of antiemetics or neuromodulators (p>0.35; matching efficacy reported in **Table 1**). Patients on prokinetics had a median TSS of 24.0 (IQR 18.6-30.0) compared to median 15.7 (IQR 8.4-26.2) in matched patients (p=0.09; **Figure 1**). There were no differences in metrics or phenotypes between patients prescribed prokinetics and matched patients not on prokinetics (p>0.15).

**Figure 1:**
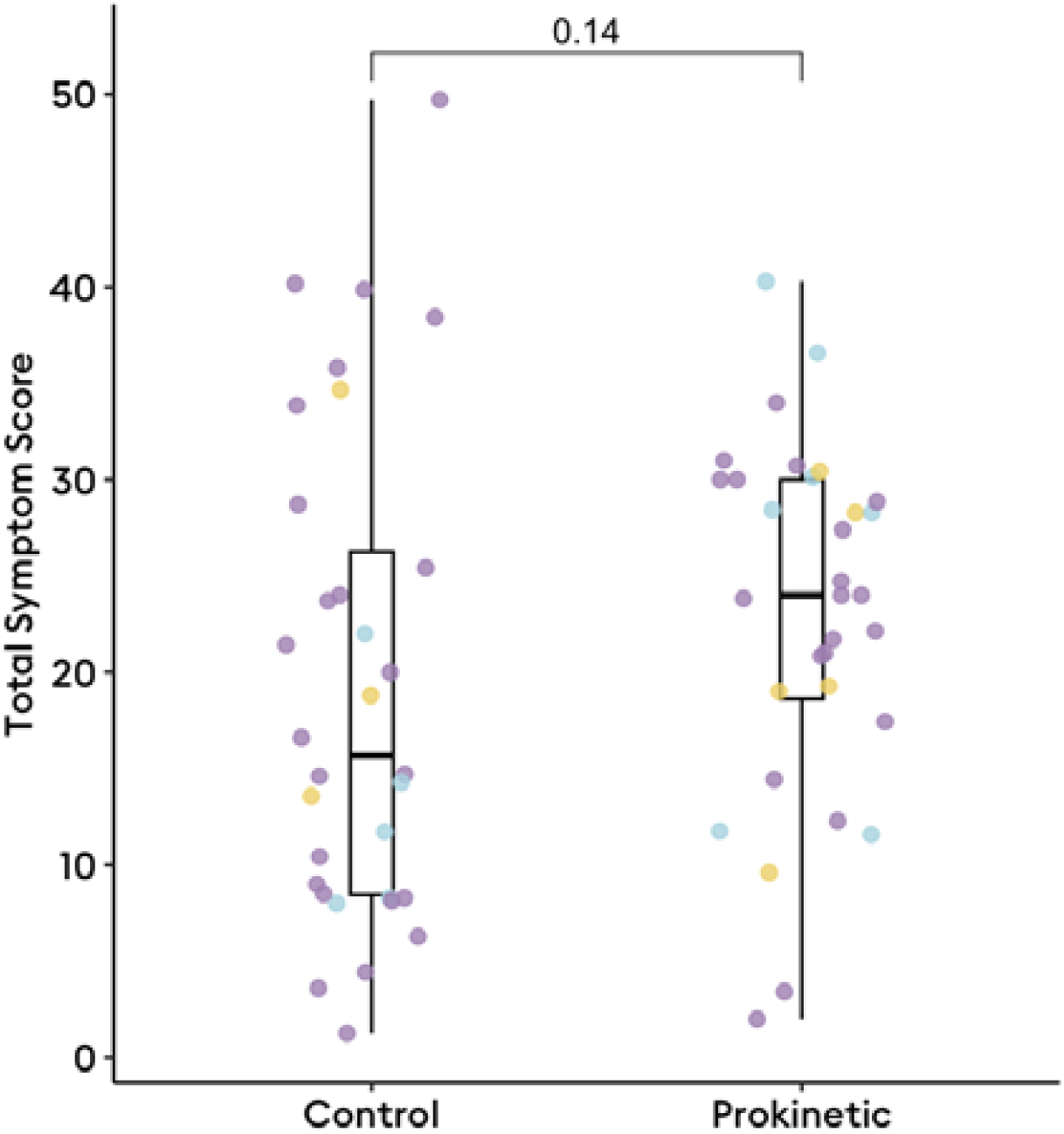
Total symptom score at 30-days after body surface gastric mapping amongst those on prokinetics (n=32) and age, sex, BMI, and medication-adjusted counterparts (n=32). Coloured dots represent Gastric Alimetry phenotypes (purple = normal phenotype; blue = neuromuscular phenotype; yellow = high-frequency phenotype).

Upon inspection of interactions between metrics and medication status, among patients taking prokinetics, lower BMI-adjusted amplitudes in the second postprandial hour were associated with lower TSS at day 30 compared to matched patients not on prokinetics (β=0.13, 95% CI 0.00-0.26, p=0.0497; **Figure 2**), such that prokinetics improve symptoms more so in patients with lower amplitudes. Similarly, lower BMI-adjusted amplitudes in the third postprandial hour was also significantly associated with lower TSS at day 30 if taking prokinetics (β=0.27, 95% CI 0.08-0.45, p=0.006; **Figure 2**) whilst this association was not seen in the matched patients not on prokinetics (β=0.20, 95% CI −0.03-0.43, p=0.09). In examining the effects of BSGM phenotype on TSS at 30 days, taking a prokinetic was not associated with symptoms in patients with normal (β=1.9, 95% CI - 4.7-8.5, p=0.6) or high-frequency phenotypes (β=-2.9, 95% CI −17-11, p=0.7); however, a neuromuscular phenotype (all had low rhythm index <0.25) was associated with significantly worse symptoms (β=12, 95% CI 0.83-23, p=0.035; **Figure 2**). This finding was further corroborated when decoupling amplitude from rhythm stability, such that prokinetics in patients with overall low rhythm index regardless of amplitude status was also associated with worse symptoms (β=15, 95% CI 4.7-25, p=0.004; **Figure 2**).

**Figure 2:**
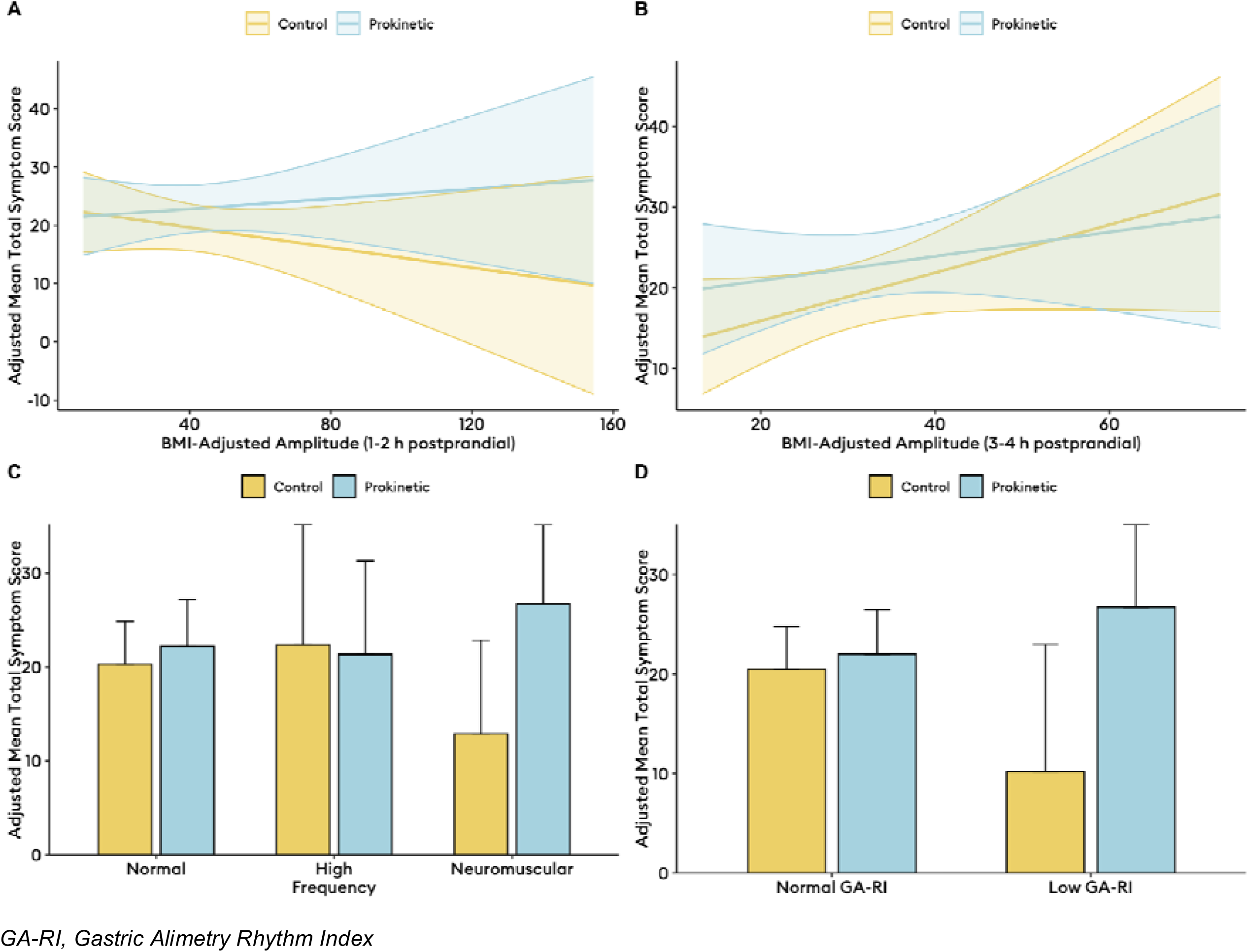
Adjusted total symptom scores calculated as marginal means from models with interaction terms for medication and Gastric Alimetry metrics (A-B) and phenotypes (C-D) in the matched cohort analysis (n=32 on prokinetics, n=32 matched symptomatic patients not on prokinetics).

### Change analysis: baseline vs on-prokinetic symptom scores

Among the 42 patients eligible for the change analysis, the baseline off-prokinetic TSS was median 18.7 (IQR 25-53), compared to median 26 (IQR 20-31; p<0.001) at follow-up of median 30 days into prokinetic therapy. Treatment responses were heterogeneous (**Figure 3**), with a MCID of 3 (mean difference in TSS −5±7) used to define treatment response. Based on the MCID, there were 8 treatment responders, and 34 non-responders, and there were no significant differences in BSGM metrics (**Figure 4**), nor Gastric Alimetry phenotypes between these groups. Of prokinetic responders, 3 (38%) had a neuromuscular phenotype, compared to 6 (17.6%) among prokinetic non-responders (p=0.3). Representative cases are shown in **Figure 5**.

**Figure 3:**
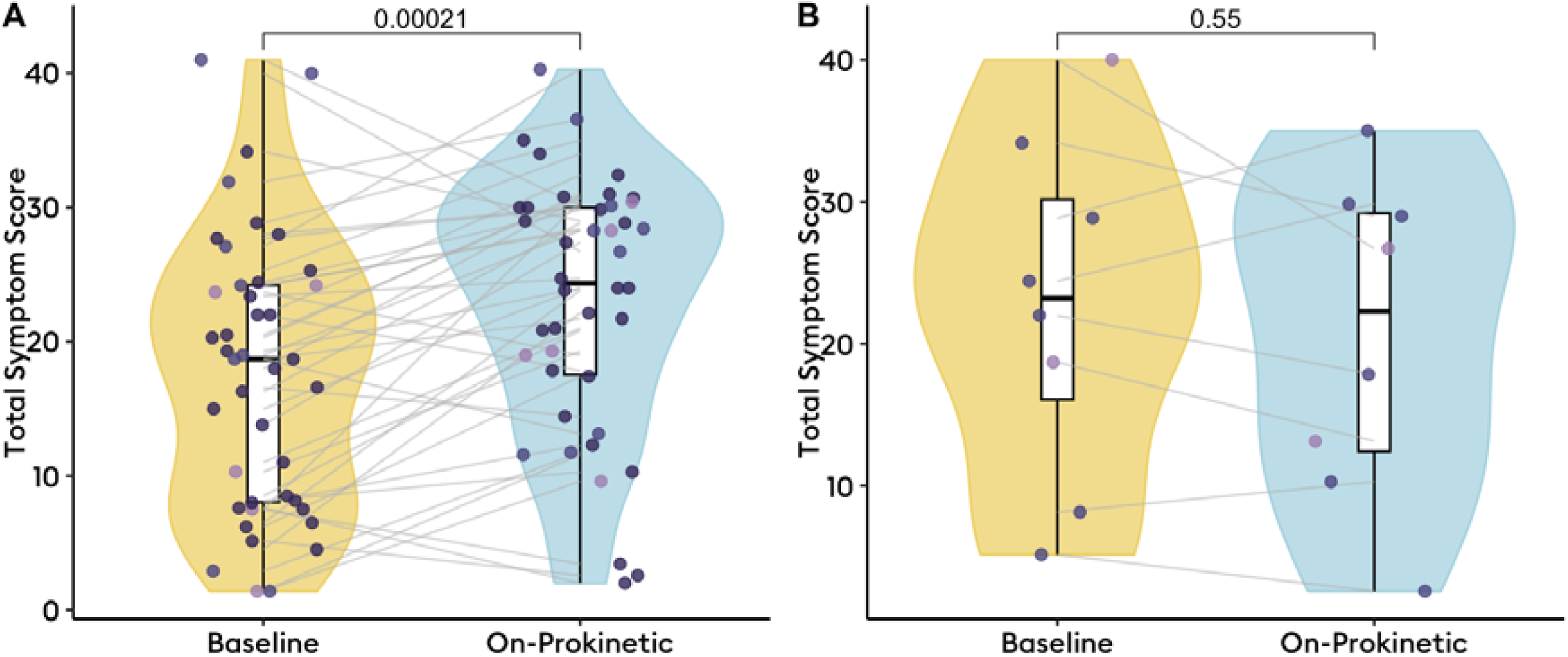
Change in total symptom score from off-prokinetic baseline to on-prokinetic time period. A: Change in symptoms upon starting prokinetic in overall cohort (n=42); B: Change in symptoms among previously prokinetic naive patients in subgroup analysis (n=8).

**Figure 4:**
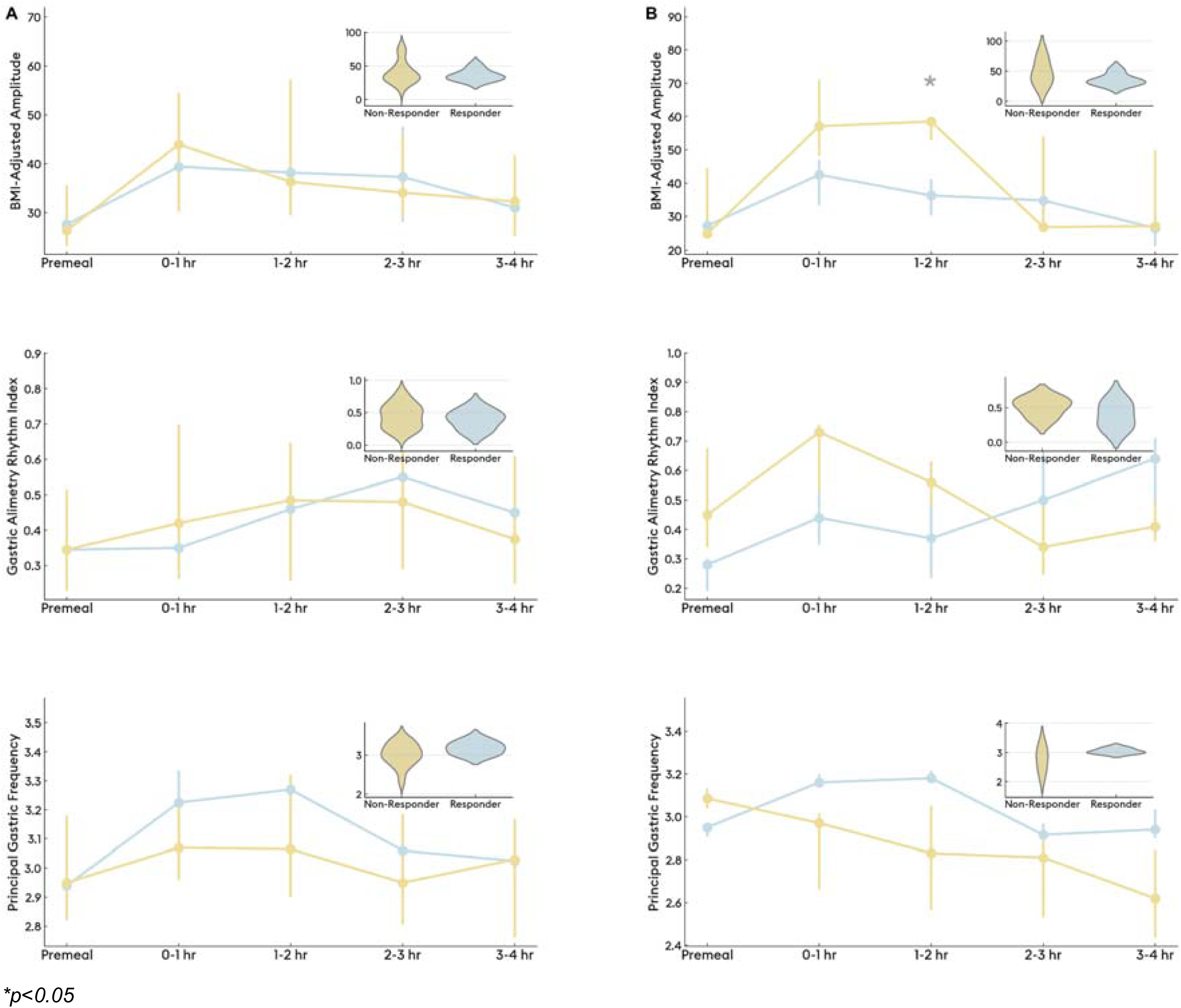
Differences in BMI-adjusted amplitude, Gastric Alimetry Rhythm Index, and Principal Gastric Frequency, between prokinetic responders and non-responders. A) Differences in BSGM metrics across the test with box plots depicting overall differences in metrics between responders and non-responders (n=42); B) Differences in BSGM metrics across the test with box plots depicting overall differences in metrics between responders and non-responders of subgroup analysis of initially prokinetic naive patients (n=8).

**Figure 5:**
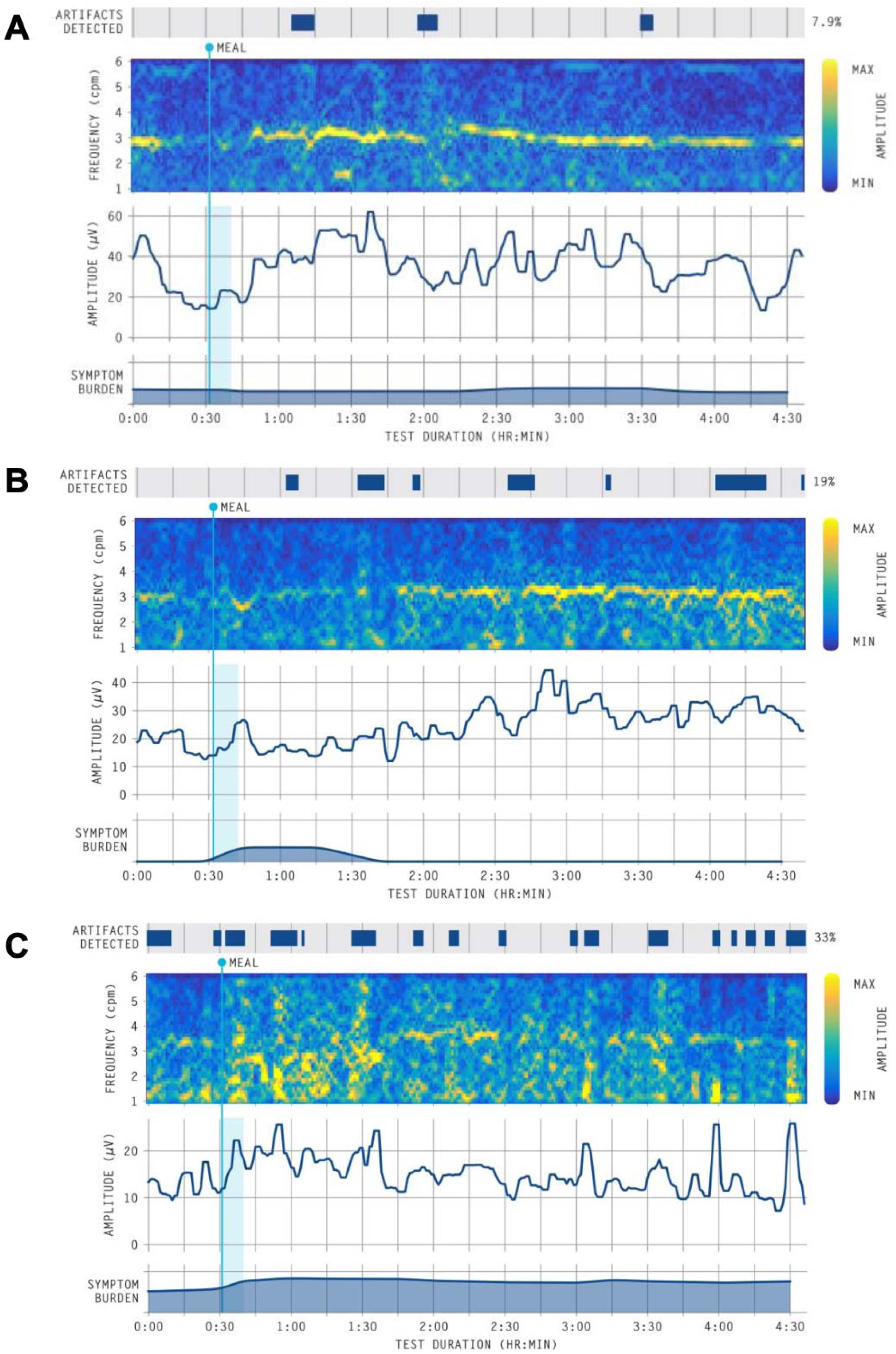
Representative spectrograms from overall cohort. A) normal spectrogram (all metrics within normative reference intervals)^16^ of patient who responded to prokinetic therapy; B) spectrogram showing a period of low-amplitude in the early postprandial phase coinciding with symptoms in a patient who responded to prokinetic therapy; C) spectrogram with unstable rhythms and high symptom burden in an individual who did not respond to prokinetic therapy.

### Subgroup analysis of prokinetic naive cohort

Among the above 42 patients, 8 were previously prokinetic naive, and started prokinetics at median 80 (IQR 34-133) days after Gastric Alimetry testing. At median 76 days into prokinetic therapy, overall median baseline TSS was 22 (IQR 12-29), which was unchanged from baseline (23 (IQR 16-30); p<0.4). In this group of previously prokinetic naive patients, the MCID was 2 (mean difference in TSS +2±6), and there were 5 prokinetic-responders (**Figure 3**). Prokinetic responders again had lower baseline BMI-adjusted amplitudes in the second postprandial hour (mean 37.5±10.6 uV in responders [n=5] vs mean 54.8±6.6 uV in non-responders [n=3], p=0.047; **Figure 4**). In this subgroup, 6/8 had a normal phenotype, and the remaining two had a neuromuscular phenotype, both represented within the prokinetic responders group (p=0.5).

## Discussion

This is the first study to demonstrate that Gastric Alimetry biomarkers may help predict response to prokinetic therapies in patients with chronic gastroduodenal symptoms. The key finding was that lower post-prandial amplitudes at baseline (i.e., not on prokinetic therapy), indicating relatively blunted meal responses, predicted benefit from prokinetics. In contrast, disrupted gastric rhythms predicted a poorer response to prokinetics. These findings were consistent across a range of analyses, suggesting a potentially important role for Gastric Alimetry assessment in guiding prokinetic therapy.

Legacy electrogastrography (EGG) studies were highly inconsistent in identifying biomarkers for treatment success in gastroduodenal disorders; with some studies suggesting abnormal myoelectrical activity predicted response to prokinetics,^18,19^ whilst others showed the opposite.^20^ Notably, legacy EGG analyses typically defined abnormalities based on duration of recording outside a broad frequency band (e.g. 2-4 cpm), and could not reliably measure amplitude due to lack of adjustment for BMI.^21^ By contrast, the refined BSGM techniques applied here identify a tight range in which the normal gastric frequency occurs (2.65-3.35 cpm) while separately defining normal rhythm index as ≥0.25 (range: 0-1), and normal amplitude as lying within the range of 22-70 μV.^16,22,23^

In applying these robust new biomarkers of gastric myoelectrical function, it is important to note that the responders in this study did not typically meet criteria for an overall ‘low amplitude phenotype’ (as defined by the BMI-adjusted amplitude being <22 μV reference range over the whole 4 hr postprandial period), which is thought to represent smooth muscle myopathy or neuromuscular disorders.^12,16,24^ Instead, responders showed weaker meal responses than non-responders, indicated by relatively lower amplitudes in the first 2 post-prandial hours (typically suppressed into the 20-40 μV range). This finding is promising for potentially identifying a broad range of patients who stand to benefit from prokinetics, being consistent with these agents’ mechanism of action, which aims to increase contraction amplitude in order to address post-prandial distress.

The importance of rhythm stability for determining the direction of symptom response in this study suggests an intact neuromuscular apparatus is required for prokinetic effects. When gastric rhythms were unstable, likely implicating deficits in the number or function of interstitial cells of Cajal,^25–27^ symptom responses were worse on prokinetics. Rhythm instability may therefore be a key biomarker of medication-refractory pathology, where escalation of therapies may be warranted. This observation is supported by two other recent studies, one in pediatric patients and another in patients on parenteral nutrition, where poor rhythm stability indicated more severe or refractory patient groups.^28,29^

This study has several limitations, primarily that it is an observational prospective cohort study.^13^ As such, linear modeling approaches were required to accommodate the real world heterogenous prokinetic prescription timings in this cohort. This work was also limited by the variety of prokinetic agents prescribed in a real world observational cohort, preventing analysis of the effects of specific prokinetics in subgroups. However it is notable that consistent statistically significant findings were still evident despite the current sample size on differential analyses, providing confidence in the data. These promising results therefore clearly motivate a multicentre prospective study to further define and expand these findings in a larger cohort, and this study is now being planned.

Previous reports using legacy EGG suggest that prokinetics increase postprandial myoelectrical power.^20,30,31^ However, these measurements were often imprecise due to noise contamination, and the use of metrics that conflated rhythm and frequency.^21^ Among several important developments in modern BSGM were to separate rhythm and frequency metrics,^16^ adjust the amplitude for BMI to account for signal dissipation with abdominal adiposity, and rigorously optimise noise and artifact rejection.^32^ In this study we applied these improved techniques, but did not specifically assess gastric myoelectrical responses before and after prokinetic initiation. This presents a further valuable direction for future physiological research, in order to determine whether legacy EGG studies showing modified EGG power are now replicated using BSGM.

Prescribing prokinetics is not without risk of adverse events, and as such patient selection for prokinetic therapy should be determined based on maximal likelihood of symptomatic benefit. This study shows Gastric Alimetry biomarkers can potentially guide selection for prokinetic therapy in patients with chronic gastroduodenal symptoms, favouring patients with stable rhythms and relatively blunted meal responses (lower amplitude myoelectrical activity in the post-prandial period). Given the importance of prokinetic prescribing globally, which is currently largely based on trial and error, further validation of these findings in a large prospective study should be prioritised.

## Data Availability

Patient data are not available.

